# Global access to handwashing: implications for COVID-19 control in low-income countries

**DOI:** 10.1101/2020.04.07.20057117

**Authors:** Michael Brauer, Jeff T Zhao, Fiona B Bennitt, Jeffrey D Stanaway

**Affiliations:** Institute for Health Metrics and Evaluation, University of Washington, Seattle WA USA; School of Population and Public Health, The University of British Columbia, Vancouver BC Canada

## Abstract

**Background:** Low-income countries have reduced health care system capacity and are therefore at risk of substantially higher COVID-19 case fatality rates than those currently seen in high-income countries. Handwashing is a key component of guidance to reduce transmission of the SARS-CoV-2 virus, responsible for the COVID-19 pandemic. Prior systematic reviews have indicated the effectiveness of handwashing to reduce transmission of respiratory viruses. In low-income countries, reduction of transmission is of paramount importance but social distancing is challenged by high population densities and access to handwashing facilities with soap and water is limited.

**Objectives:** To estimate global access to handwashing with soap and water to inform use of handwashing in the prevention of COVID-19 transmission.

**Methods:** We utilized observational surveys and spatiotemporal Gaussian process regression modeling in the context of the Global Burden of Diseases, Injuries, and Risk Factors Study, to estimate access to a handwashing station with available soap and water for 1062 locations from 1990 to 2019.

**Results:** Despite overall improvements from 1990 (33.6% [95% uncertainty interval 31.5–35.6] without access) to 2019, globally in 2019, 2.02 (1.91–2.14) billion people—26.1% (24.7–27.7) of the global population lacked access to handwashing with available soap and water. More than 50% of the population in sub-Saharan Africa and Oceania were without access to handwashing in 2019, while in eight countries, more 50 million or more persons lacked access.

**Discussion:** For populations without handwashing access, immediate improvements in access or alternative strategies are urgently needed, while disparities in handwashing access should be incorporated into COVID-19 forecasting models when applied to low-income countries.

**Funding:** Bill & Melinda Gates Foundation. MB was supported in part by the *Pathways to Equitable Healthy Cities* grant from the Wellcome Trust.

## Introduction

In the initial months of the Coronavirus Disease 2019 (COVID-19) pandemic, the outbreak has been concentrated in middle- and high-income countries: initially China, followed by high-income east Asia, the Middle East, Europe, and North America. Even as health care systems in these relatively well-equipped regions are strained, there will soon be a need to focus on populations in low-income countries (LICs) where health care resources are limited, even before facing the demands of the pandemic. Along with social distancing, handwashing has been advised repeatedly as one of the key actions to reduce transmission of the SARS-CoV-2 virus, responsible for the COVID-19 pandemic (Centers for Disease Control and Prevention; World Health Organization, 2020). In a systematic review of physical interventions employed to reduce the transmission of respiratory viruses, handwashing was indicated to be effective with a meta-analytic summary estimate of a 45-55% reduction in transmission(Jefferson et al. 2009, 2011). Similarly, a systematic review of the effectiveness of personal protective measures in preventing H1N1 pandemic influenza transmission in human populations indicated a 38% reduction in transmission with handwashing, while mask use was less effective and data for cough etiquette insufficient (Saunders-Hastings et al. 2017). A review of influenza transmission in adults also concluded that handwashing was effective in reducing transmission (Smith et al. 2015).

While access to handwashing facilities with soap and water is near-universal in high-income countries, the same is not true for LICs. As limited access to handwashing facilities may promote the spread and magnitude of the COVID-19 pandemic in low-income countries, governments and aid agencies may prioritise rapid deployment of access or alternatives such as alcohol-containing handrub solutions to those locations without access. Further, application of COVID-19 forecasting models, especially those derived from high- and middle-income country data, to LICs may need to account for handwashing access.

In the context of the Global Burden of Diseases, Injuries, and Risk Factors Study (GBD), we estimated access to a handwashing station with available soap and water for 1062 locations from 1990 to 2019.

## Methods

Access to a handwashing station with available soap and water was based on the WHO/UNICEF Joint Monitoring Programme for Water Supply, Sanitation and Hygiene (The WHO/UNICEF Joint Monitoring Programme for Water Supply, Sanitation and Hygiene (JMP)) definition for basic hygiene of “Availability of a handwashing facility on premises with soap and water,” where a handwashing facility was defined according to Sustainable Development Goal 6.2.1 as “a device to contain, transport or regulate the flow of water to facilitate handwashing.” (Progress on Drinking Water, Sanitation and Hygiene Update and SDG Baselines 2017 Launch version July 12 Main report 2017). This measure is a proxy of actual handwashing practice, but one with improved accuracy compared with other proxy measures such as self-reported handwashing (Ram 2013). A full list of data sources (Excel Table S1) as well as a tabulation of sources by country and year (Excel Table S2) can be found in the Supplemental Material. We systematically reviewed and re-extracted all primary input sources to ensure data quality. When surveys were extracted at the household level, the household weights were multiplied by the household size to provide individual weights for tabulating access by individuals. Thus, all estimates are provided at the individual level to ensure larger households are properly captured. All data were extracted from the original microdata at either the household or individual level, processed to standardize indicators and expanded household weights to individual weights, and then tabulated by national or subnational location. We excluded sources that did not include sample weights, were not representative of the population of a GBD (national or subnational) location, were missing data for handwashing station, available water, and/or available soap, or reported a single, aggregated “handwashing agent” indicator that included ash, sand, or soil in addition to soap. Country-specific Demographic and Health Surveys (DHS), Multiple Indicator Cluster Surveys (MICS), and Performance Monitoring and Accountability 2020 (PMA2020) surveys as well as several censuses conducted from 2008 to 2019 were included as inputs. Input data sources prior to 2008 were excluded due to data quality with respect to capturing all three aspects satisfying the definition of basic hygiene: 1) presence of a station, with available 2) soap and 3) water.

These data sources were used as inputs into the GBD 2017 spatiotemporal Gaussian process regression (ST-GPR) modelling tool, which has been described in detail elsewhere (Stanaway et al. 2018). Briefly, this tool uses a two-stage modelling framework that includes a mixed effect linear regression model, followed by a ST-GPR step to model the proportion of the population with access to a handwashing station on premises with available soap and water by year and location from 1990 – 2019. This model allows us to borrow strength across space and time, and leverage information from predictive covariates to produce a complete set of estimates for all GBD locations and estimation years. The linear regression model included as covariates the proportion of individuals with access to piped water (modeled in a similar fashion from country-specific surveys) and the Socio-demographic Index (SDI, a composite measure including income per capita, education, and fertility), with region and super-region random effects. In total 153 surveys from 88 countries and territories were included as inputs (Figure 1). Piped water was included as a covariate as there was an observed statistical relationship between piped water and handwashing access in surveys where both were collected, and since more complete information on piped water was available from surveys than for handwashing access. We therefore use estimates of piped water as a predictive covariate to improve estimates of handwashing. All of the included data sources were from the years 2008–2019. In total we estimated access for 204 national and non-sovereign locations (eg, Guam, Puerto Rico) and 858 sub-national administrative areas. In India, we also model urban and rural locations as source input data and covariates have been coded to urban and rural locations through the GBD collaborator network. In several other locations, sub-national administrative boundaries define urban (metropolitan) areas allowing for some inferences on access to handwashing facilities with available soap and water.

**Figure 1.**
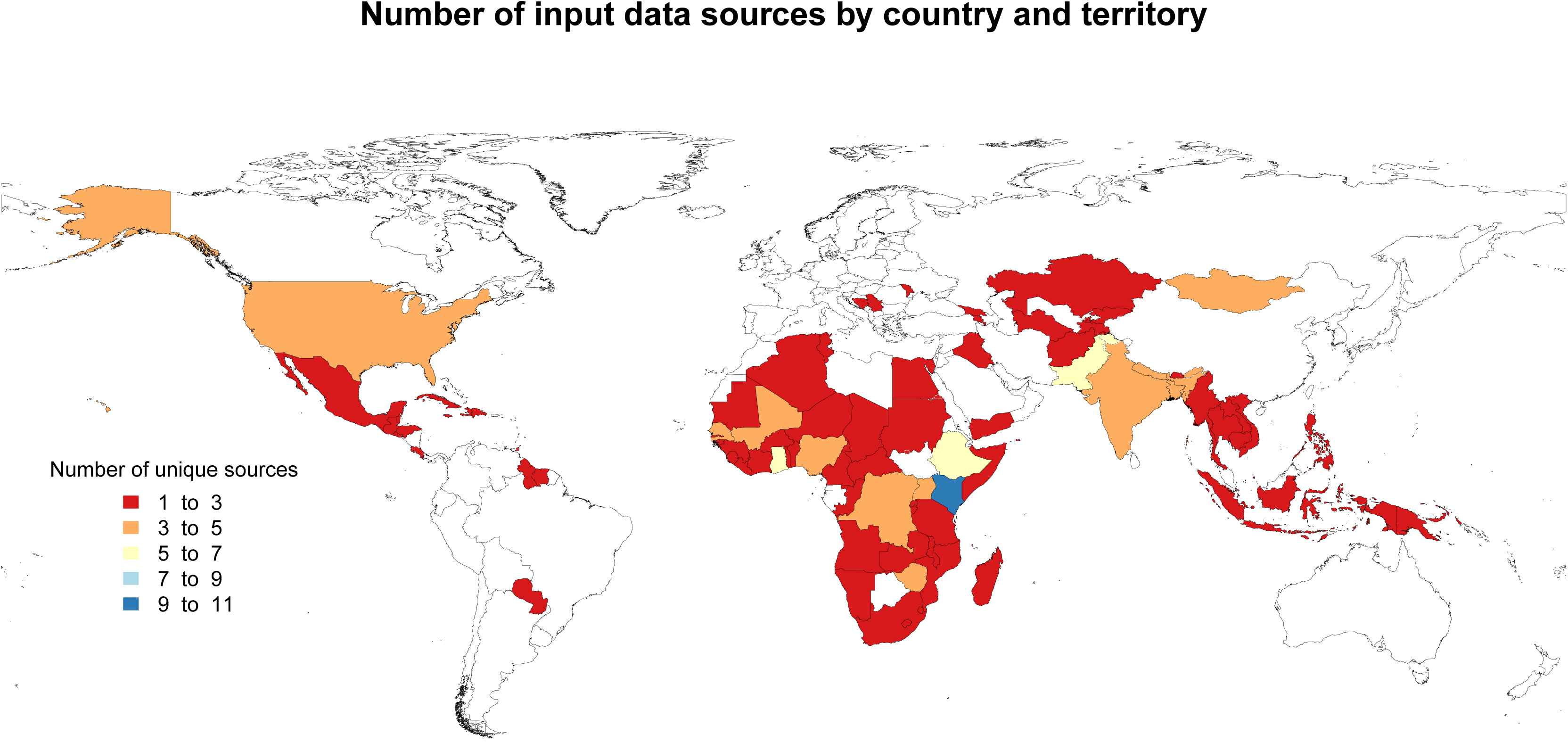
Number of input data sources by country and territory, 2019. See Excel Table S1 for input data sources and Excel Table S2 for corresponding numeric data.

## Results

In 2019, 2.02 (95% uncertainty interval [UI] 1.91–2.14) billion people—26.1% of the global population—were estimated to lack access to handwashing facilities with soap and water. We estimated higher proportions without access in LICs, especially those in sub-Saharan Africa, south Asia and the Caribbean (Figure 2). In 46 countries more than half of the population lacked access, and in eight countries (India, Nigeria, China, Ethiopia, Democratic Republic of the Congo, Bangladesh, Pakistan, Indonesia) more than 50 million persons were estimated to be without handwashing access. Example figures showing the STGPR estimates and fits to survey data for these locations are provided in the Supplemental Material (Figures S1-S8). In India alone, some 499 million (393–608) people lacked access. Estimates for all national (Excel File S3) and sub-national (Excel File S4) locations for all years from 1990 – 2019 are provided in the Supplemental Material.

**Figure 2.**
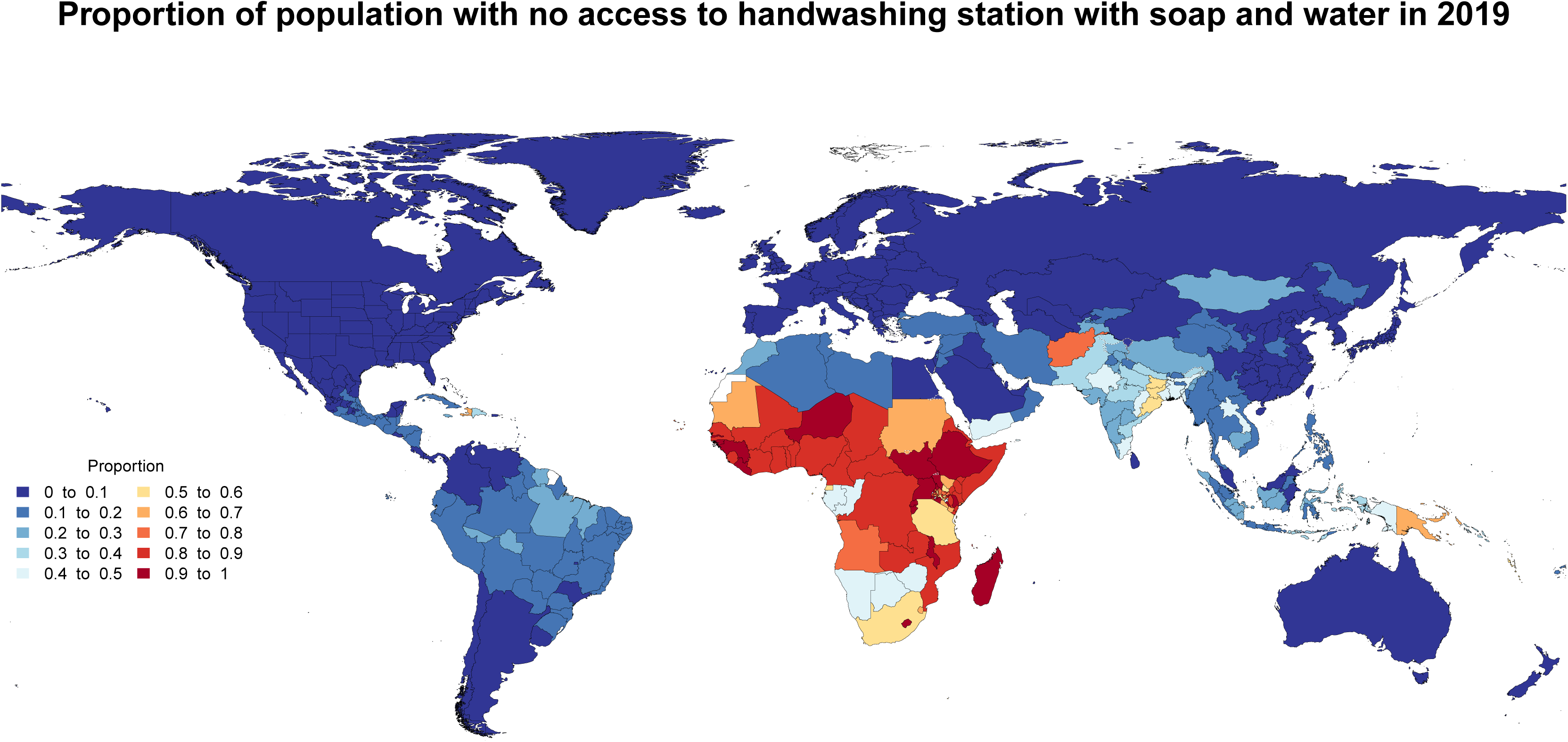
Estimated Proportion of the population with no access to a handwashing station with soap and water in 2019. See Excel Tables S3 and S4 for corresponding numeric data for 2019 and all years (1990–2019).

While the numbers of people lacking access to handwashing facilities with soap and water are large, there have been substantial improvements in many regions of the world (Table 1). In particular, we estimated that there were more than 25% reductions in lack of access from 1990 to 2019 in 17 countries and territories (Paraguay, Gabon, Bhutan, Equatorial Guinea, Oman, Botswana, São Tomé and Príncipe, Tanzania, Palestine, Guatemala, Nepal, Marshall Islands, Morocco, Tokelau, Tuvalu, Congo, Saudi Arabia) (Excel File S3, Figures S1 – S21). These reductions parallel region-wide improvements in some cases (eg, north Africa and the Middle East, Latin America) while others are unique examples within their respective regions (eg, Bhutan, Gabon, Equatorial Guinea, Tanzania, Botswana, São Tomé and Príncipe). Overall however, little progress has been made throughout most of sub-Saharan Africa. Access is strongly related to SDI although substantial variability is present at similar SDI levels and within regions (Figure 3).

**Table 1.** Estimated Percentage of population without access to handwashing with soap and water globally and by GBD region, 1990 and 2019. GBD=Global Burden of Diseases, Injuries, and Risk Factors Study. UI=uncertainty interval. Estimates were based upon input survey data inputs into a spatiotemporal Gaussian process regression model with access to piped water and the Socio-demographic Index as model covariates.

| Region | Percentage of population without access, 1990 (95% UI) | Percentage of population without access, 2019 (95% UI) |
| --- | --- | --- |
| Global | 33.6 ( 31.5 , 35.6 ) | 26.1 ( 24.7 , 27.7 ) |
| East Asia | 26.1 ( 24.2 , 28.1 ) | 7.7 ( 6.9 , 8.4 ) |
| Southeast Asia | 28.6 ( 24.1 , 33.5 ) | 16.6 ( 14.1 , 19.5 ) |
| Oceania | 63.3 ( 56.0 , 69.4 ) | 56.6 ( 52.5 , 60.5 ) |
| Central Asia | 13.6 ( 11.3 , 16.4 ) | 7.8 ( 6.5 , 9.4 ) |
| Central Europe | 5.5 ( 4.5 , 6.9 ) | 2.8 ( 2.3 , 3.4 ) |
| Eastern Europe | 6.4 ( 4.6 , 8.5 ) | 3.9 ( 2.7 , 5.3 ) |
| High-income Asia Pacific | 1.4 ( 1.0 , 2.0 ) | 1.0 ( 0.6 , 1.4 ) |
| Australasia | 1.5 ( 0.9 , 2.1 ) | 1.0 ( 0.7 , 1.6 ) |
| Western Europe | 1.3 ( 1.1 , 1.6 ) | 0.9 ( 0.8 , 1.1 ) |
| Southern Latin America | 5.4 ( 3.6 , 7.9 ) | 1.5 ( 1.0 , 2.1 ) |
| High-income North America | 0.6 ( 0.4 , 0.9 ) | 0.4 ( 0.3 , 0.5 ) |
| Caribbean | 35.4 ( 31.6 , 39.5 ) | 34.1 ( 31.1 , 36.8 ) |
| Andean Latin America | 23.8 ( 18.5 , 29.8 ) | 12.9 ( 9.6 , 16.8 ) |
| Central Latin America | 18.6 ( 15.2 , 23.0 ) | 10.6 ( 8.8 , 12.6 ) |
| Tropical Latin America | 29.9 ( 21.0 , 40.5 ) | 13.8 ( 9.1 , 20.5 ) |
| North Africa and Middle East | 33.0 ( 29.9 , 36.3 ) | 21.1 ( 19.8 , 22.6 ) |
| South Asia | 59.5 ( 50.3 , 68.7 ) | 37.2 ( 31.2 , 43.5 ) |
| Central sub-Saharan Africa | 89.7 ( 86.9 , 92.0 ) | 81.7 ( 79.1 , 84.1 ) |
| Eastern sub-Saharan Africa | 92.4 ( 90.9 , 93.6 ) | 83.6 ( 82.3 , 84.9 ) |
| Southern sub-Saharan Africa | 64.8 ( 56.1 , 72.6 ) | 51.2 ( 45.7 , 56.7 ) |
| Western sub-Saharan Africa | 90.7 ( 88.4 , 92.7 ) | 85.5 ( 83.5 , 87.3 ) |

**Figure 3.**
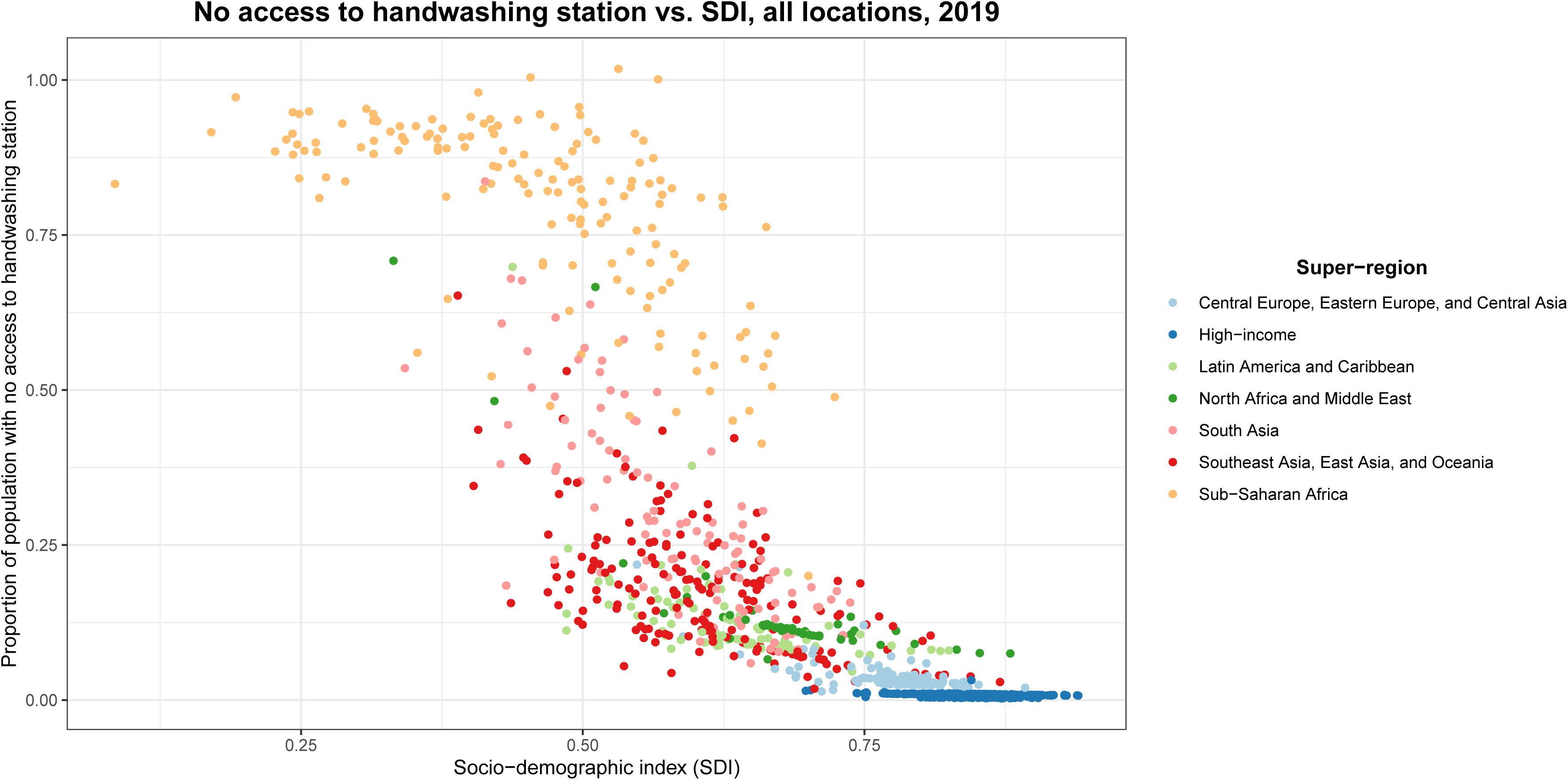
Estimated relationship between no access to a handwashing station with soap and water and SDI, by GBD region, 2019. SDI=Socio-demographic Index.

## Discussion

Low-income countries have reduced health care system capacity and are therefore at risk of substantially higher COVID-19 case fatality rates than those currently seen in high-income countries. In this context, suppression of transmission has heightened urgency. Inadequate access to handwashing facilities with soap and water remains prevalent in many LICs, and this is likely to facilitate COVID-19 transmission. Rural populations have disproportionately poor access to handwashing facilities; for example, across India, state-level estimates ranged from 6% without access (Mizoram) to 31% (Odisha) in urban areas, and from 13% without access (rural Delhi) to 68% (Jharkhand) in rural areas. However, access is also limited in urban slums and other informal settlements. Our estimates indicate that, in 2019, 15% of the population in urban Delhi, 58% in Addis Ababa (province) and 54% in Nairobi (province) lacked handwashing access. Should the pandemic coincide with water shortages, such as those seen in Cape Town and Chennai in 2019 (Holden and Doshi 2019), access will be further restricted and will disproportionately harm those who can least afford to pay for water. In densely-populated urban areas, social distancing is also very challenging given high population densities. Further, effective within-household quarantine is likely impossible with larger families living together in a single home. Reducing COVID-19 transmission in high-density urban areas with low access to handwashing may prove especially difficult and will require urgent attention and implementation of alternative strategies to those implemented in high- and middle-income countries.

Alcohol-containing handrub solutions are an efficacious alternative to hand washing with soap and water (Kampf 2018). Guidance exists for local production of handrub solutions, and evidence from prior epidemics supports the effectiveness of local production (World Health Organization 2010). However, reliance on handrub solutions is less desirable compared to handwashing given requirements for sustainable production and distribution, cost implications for low-income populations, concerns regarding flammability of reagents, and the potential for poisoning due to ingestion (Gormley et al. 2012). Further, access to handwashing with soap and water can offer more equitable and lasting protection in future epidemics and also protect against non-epidemic transmission of diarrheal disease (Wolf et al. 2018) and lower respiratory infections (Mbakaya et al. 2017; Rabie and Curtis 2006) if sufficiently maintained. Indeed, in 2017 inadequate access to handwashing was estimated to be sufficiently maintained. Indeed, in 2017 inadequate access to handwashing was estimated to be responsible for 35% of the global diarrheal disease burden and 9.7% of the global burden from lower respiratory infections, in total accounting for 38.4 million DALYs (95% UI 22.8–52.0), and 707 000 deaths (416 000–1 022 000) (Stanaway et al. 2018). Increases in these and other common causes of death unrelated to COVID-19 are also likely to be affected as LIC health care systems are overwhelmed during the pandemic. Immediate efforts to increase handwashing access or alternatives could help alleviate some of the baseline disease burden from diarrhea and lower respiratory infections during the pandemic and spare valuable resources to focus on COVID-19 cases.

Progress towards improving access to handwashing has been accelerated given the recognition of its importance in two of the Millennium Development Goals, *reducing childhood mortality* and *combatting HIV/AIDS, malaria, and other diseases*, and more recently as part of Sustainable Development Goal 6*: to ensure availability and sustainable management of water and sanitation for all*, with the indicator 6.2.1 used in this analysis (6.2.1 Proportion of Population with Basic Handwashing Facilities on Premises - SDG Global Indicator Platform). With estimates provided here for 2019, this analysis presents the most recent and comprehensive global estimates of handwashing access to date, including estimates for 204 national and non-sovereign locations and 858 sub-national administrative areas. The enhanced spatial resolution gained by including sub-national locations reduces the potential for spatial misalignment between survey responses and population density, thereby providing a more accurate picture of the true population with access in each country, and offers policy makers more detailed and actionable evidence.

As in most global-scale analyses, these estimates include several limitations. Most importantly, while we developed comprehensive global estimates, a number of potential sources from 2000 to 2005 reported a soap indicator which included “other handwashing agents” such as ash, sand, or soil and therefore did not meet JMP’s definition for basic hygiene (The WHO/UNICEF Joint Monitoring Programme for Water Supply, Sanitation and Hygiene (JMP)). This considerable limitation is reflected in the uncertainty in national-level estimates. These national level estimates are based on recently available survey data; however, the year of the most recent survey varies by location, also contributing to uncertainty. While our spatiotemporal model estimates incorporate trends to estimate access in 2019, the most recent source input data may have been from earlier years (Excel File S1, Excel File S2). Similarly, estimates for years prior to 2008 were derived from the ST-GPR model and not from survey inputs. This is reflected in uncertainty intervals (Figures S22-S29). Further, our estimates likely do not reflect recent disruptive events such as conflict, large-scale migration, or natural disasters. These events are likely to increase the number of people living without access to handwashing and also to increase the number of people living in densely populated settings where social distancing is also challenged. As the input data sources are based on snapshot observations, they also do not reflect intermittent or discontinued water supplies which can be common in many locations. Data coverage was best throughout Africa and relatively poor in South America and south and east Asia (Figure 1). Our estimates also do not include access to handwashing facilities in non-household settings such as schools, workplaces health care facilities and other public locations such as markets. The extent to which these locations may remain accessible during stay at home and other restrictions related to the COVID-19 response is unknown.

Our estimates update and expand upon other available estimates of handwashing access developed by the WHO and the WHO/UNICEF Joint Monitoring Program (JMP). Specifically, we produce estimates on a much broader spatial and temporal scale. The most recent JMP report includes estimates for 96 countries up to the year 2017 (Progress on household drinking water, sanitation and hygiene | 2000–2017: Special focus on inequalities 2019), as well as urban and rural estimates for most of those locations, while we estimate for 1062 national and subnational locations from 1990–2019. We are able to do so because our modeling methods borrow strength across time and space and leverage information from predictive covariates to produce estimates for locations without data. The first stage of our modeling process involves running a mixed-effects linear model, with fixed effects on socio-demographic index (SDI) and proportion of the population with access to a piped water source, and country- and region-level random effects. These covariates inform the model fit in all locations, especially those without data. After the linear model is run, the next steps involve spatial and temporal smoothing. Essentially, when estimating for any given location-year, data points that are geographically and temporally proximate are weighted more heavily than more distant data points. For example, if a subnational location has no data, the national-level data informs the model fit; similarly, in a country with no data, the region-level data informs its estimates. Our estimation models allow for non-linear trends, borrows strength from similar sources to extend spatial coverage, uses predictive covariates to inform estimates, and leads to more robust estimates by including both large and smaller surveys. In comparison, JMP uses a set of rules to linearly interpolate and extrapolate data separately for each location.

JMP reports three different levels of access to handwashing: “basic”, meaning access to a handwashing facility with soap and water; “limited”, a facility with no soap or water, and “none.” Because we defined anyone without access to a handwashing facility with soap and water as having no access, we compared our estimates to the sum of JMP’s “limited” category (access to a facility, but no soap or water) and “no facility” category. The overall correlation coefficient between the two sets of estimates (for locations where JMP provides estimates) was 0.94, while the year-specific correlation coefficient ranged from 0.91 (comparing 2017 estimates, the most recent year of JMP estimates) to 0.985 (comparing 2003 estimates).

We have presented estimates of access to handwashing facilities which is an imperfect proxy for actual handwashing behavior, as previous research indicates that the prevalence of actual handwashing is lower than the prevalence of access to handwashing facilities (Freeman et al. 2014; Wolf et al. 2018). Despite this limitation our approach has two notable advantages. First, whereas adequate data exist to model access, data on handwashing behavior are inadequate to support a reliable global model. Second, the proportion of people with access to handwashing likely represents the upper limit of the proportion of people engaged in handwashing. Since concerns about COVID transmission have likely increased handwashing well above regular levels, access might better represent current handwashing levels.

Understanding disparities in handwashing access must be considered in COVID-19 forecasting models, especially when applied to low-income countries. Specifically, as the pandemic has initially affected mainly high income countries with high handwashing access, transfer of transmission efficiency estimates and/or epidemic time trends from such settings should be adjusted when applied to settings with poor access to handwashing. For example, R_0_ estimates may be higher and/or more uncertain in settings with poor handwashing access compared to those estimated from high-income country settings where the first wave of the epidemic has largely been completed. Further, the impacts of social distancing which appear to have been successful in reducing transmission in high income country settings (which typically have high levels of handwashing access) may be less effective in settings where handwashing access is low. We suggest including variation in handwashing access as a covariate in the application of forecasts to data sparse low income countries.

In the context of the global impact of COVID-19, inadequate access to handwashing affects a large proportion of the world’s population and may undermine strategies for control of disease transmission. For those locations currently without access, alternative strategies are urgently needed. To the extent that access can be implemented in the short-term, opportunities exist to both help reduce COVID-19 transmission and to reduce in the long-term the 707 000 deaths from diarrheal disease and lower respiratory infections that are attributable to no handwashing access.

## Data Availability

Data referred to in the manuscript can be made available upon specific requests to the authors.

## Acknowledgments

This work was supported by funding from the Bill & Melinda Gates Foundation. MB acknowledges support from the *Pathways to Equitable Healthy Cities* grant from the Wellcome Trust.

